# *Antonyms:* A Performance Metric to Assess Word-Finding Difficulty in Multiple Sclerosis

**DOI:** 10.64898/2026.09.21.26363587

**Authors:** James F. Sumowski, Emily Dvorak

**Affiliations:** Department of Neurology, Icahn School of Medicine at Mount Sinai, New York, NY, USA; Department of Psychiatry, Weill Cornell Medical College, New York, NY, USA

**Author notes:** Corresponding Author: James F. Sumowski, PhD, Corinne Goldsmith Dickinson Center for Multiple Sclerosis, Department of Neurology, Icahn School of Medicine at Mount Sinai, 5 East 98^th^ Street, Box 1138, New York, NY, USA.

## Abstract

Word-finding difficulty (tip-of-the-tongue phenomenon) is among the most prevalent changes reported by persons with multiple sclerosis (MS), but we lack validated performance metrics to assess this common symptom. *Antonyms* is a novel task evaluating retrieval of target phonological codes *via* a brief, clinically-feasible paradigm that controls for other cognitive processes. Test-retest reliability was good (0.85). Among 660 patients, *Antonyms* demonstrated better criterion validity than traditional language tasks (e.g., animal generation, Boston Naming Test). Demonstrating discriminate validity, *Antonyms* was specifically related to patient-reported difficulties with expressive language rather than working memory, executive/speed, or episodic memory. The *Antonyms* task is provided herein.

## INTRODUCTION

Word-finding difficulty is among the most prevalent and earliest cognitive changes reported by persons with multiple sclerosis (MS),^1–3^ but there is little research on word-finding in MS, in part because we lack validated metrics sensitive to MS-related word-finding difficulty. Traditional neuropsychological language tasks (e.g., Boston Naming Test [BNT], rapid word generation) were developed for aphasia and appear insensitive to subtle but impactful MS-related changes.^2^

Successful word finding requires rapid retrieval of the semantic content related to a word and the phonological codes representing that target word.^4–5^ Persons with MS describe a tip-of-the-tongue phenomenon during which they know what they want to express (semantic meaning), but they cannot retrieve the word (phonological code). Implication of phonological code retrieval aligns with our observation that phonemic processing is weaker in MS and linked to worse patient-reported word-finding difficulty.^6^

We developed a novel paradigm requiring rapid retrieval of specific target words cued by over-learned associations (i.e., common antonyms), which reduces semantic processing to better capture individual differences in phonological code retrieval. A control task further isolates code retrieval from other input and output demands, including articulation. Herein we describe task development and provide data on reliability and validity of this approach.

## METHODS

### Development of the *Antonyms* Task

Several words have over-learned opposites (e.g., in–out, up–down); presenting part of the antonym pair (cue word) strongly activates the other.^7^ For example, if asked for the first word that comes to mind when presented with “hot,” “good,” and “true,” the most common responses are “cold,” “bad,” and “false.” We developed a paradigm during which persons are presented with cue words of well-known antonym pairs (e.g., “young – _______”) and instructed to quickly read each cue aloud (“young”) and state its opposite aloud (“old”). The pilot task consisted of 50 items (two columns of 25) to be completed in rapid succession. As a control procedure, persons were subsequently presented with the 50 cues paired with answers (e.g., “young – old”) to be read aloud in rapid succession. The *Antonyms* score is derived by subtracting control time from antonym retrieval time to isolate phonological code retrieval (word-finding) latency from reading speed. Time to completion was the dependent variable because high accuracy was expected by design. To optimize this approach, the 50-item pilot task was completed by 170 native English speakers with MS (**Table S1**), from which 10 items with accuracy <90% or multiple meanings (e.g., “soft”) were removed to derive the final 40-item *Antonyms* task (**see Appendix**). Test-retest reliability (ICC[95%CI]; two-way mixed, absolute agreement) of the final 40-item *Antonyms* task was good (0.85[0.79, 0.90]) among 102 patients with MS (**Figure S1**), which exceeded test-retest reliabilities of traditional verbal fluency tasks assessed concurrently in the same sample (Category Fluency 0.64[0.51, 0.74]; Letter Fluency 0.61[0.48, 0.72]).

### Evaluation of Task Validity

#### Samples

Criterion validity and discriminant validity of *Antonyms* were assessed in samples of native English speakers with MS aged 18 to 59 years without another neurologic condition or serious mental illness (e.g., schizophrenia) who completed (a) the year-six follow-up of the Reserve against Disability in Early MS (RADIEMS) cohort study (research sample; IRB-approved, participants provided written informed consent) or (b) standard-of-care cognitive screenings at our MS center between November 2023 and June 2026 (clinical sample; *Antonyms* added to core clinical battery in November 2023; data captured via IRB-approved chart review).

#### Criterion Validity

##### Procedures

All patients reported frequency of “having a word ‘on the tip of your tongue’ but with difficulty getting it out” as never, rarely, sometimes, fairly often, or very often; to derive more robust sample sizes representing each level, ratings were collapsed into “never/rarely,” “sometimes,” and “often.” Both samples completed *Antonyms* and the traditional verbal fluency task requiring rapid generation of animal names (Category Fluency). The research sample also generated as many words as possible starting with the letter “f” (Letter Fluency); the clinical sample was asked to retrieve names of visually-presented drawings of low frequency objects (BNT). For comparison, both samples completed the Symbol Digits Modalities Test (SDMT, oral) as a high-sensitivity, non-specific screener of overall MS-related cognitive changes.^8^

##### Statistical Analyses

Metrics were evaluated for outliers (±3.0*IQR), skewness (≥±0.8), and kurtosis (≥±1.0), and were transformed appropriately. For the research sample, MANCOVAs investigated performance differences on *Antonyms*, Category Fluency, Letter Fluency, and SDMT across levels of patient-reported word-finding difficulty, first adjusting for age, sex, and education, then additionally adjusting for premorbid verbal ability (Wechsler Test of Adult Reading, WTAR), and then also adjusting for depression symptoms (Beck Depression Inventory, Fast Screen; performed because depression may mediate associations between subjective and objective cognitive functions, especially on timed tasks.^9^) For the clinical sample, the same MANCOVAs investigated differences in *Antonyms*, Category Fluency, BNT, and SDMT across levels of patient-reported word-finding (depression assessed with Hospital Anxiety and Depression Scale, Depression Subscale). Finally, combining samples for greater statistical power, the same MANCOVAs investigated differences in *Antonyms*, Category Fluency, and SDMT across levels of patient-reported word-finding difficulty (models additionally adjusted for sample; depression normalized 0 to 1 within each sample to account for different scales).

#### Discriminant Validity

In both samples, the Multiple Sclerosis Cognitive Scale (MSCS)^10^ assessed patient-reported cognitive difficulty across subscales of Expressive Language, Working Memory, Episodic Memory, and Executive/Speed. Pearson partial correlations investigated associations between *Antonyms* and MSCS subscales, first adjusting for age, sex, education, and sample, then additionally adjusting for premorbid verbal ability, and then also adjusting for depression symptoms. Steiger’s Z-tests (two-tailed) examined if *Antonyms* was significantly more related to patient-reported difficulty in Expressive Language than to other subscales.

## RESULTS

### Samples

See **Table 1** for characteristics of the Research (n=152) and Clinical (n=508) samples.

**Table 1.** Sample Characteristics. Demographic and clinical characteristics are shown for the research sample, clinical sample, and the combined sample. Premorbid verbal ability was estimated with the Wechsler Test of Adult Reading (WTAR) proxy of WAIS Verbal Comprehension Index (VCI).

| **Characteristic** | **Research Sample** | **Clinical Sample** | **Combined Sample** |
| --- | --- | --- | --- |
| Age, years, mean (sd) | 41.2 (7.7) | 42.3 (10.1) | 42.0 (9.6) |
| Sex, N (%) |  |  |  |
| Women | 104 (68.4) | 383 (75.4) | 487 (73.8) |
| Men | 48 (31.6) | 125 (24.6) | 173 (26.2) |
| Race and Ethnicity, N (%) |  |  |  |
| Asian | 6 (3.9) | 17 (3.3) | 23 (3.5) |
| Black, non-Latino/a | 25 (16.4) | 96 (18.9) | 121 (18.3) |
| Latino/a | 38 (25.0) | 75 (14.8) | 113 (17.1) |
| White, non-Latino/a | 83 (54.6) | 320 (63.0) | 403 (61.1) |
| Progressive Disease Course, N (%) |  |  |  |
| No | 152 (100) | 465 (91.5) | 617 (93.5) |
| Yes | 0 (0) | 43 (8.5) | 43 (6.5) |
| Years since diagnosis, median (IQR) | 8.6 (7.3, 9.9) | 6.0 [2.0, 13.0] | 7.7 [3.0, 11.6] |
| Bachelor's degree, N (%) | 125 (82.2) | 398 (78.3) | 523 (79.2) |
| Premorbid Verbal Ability, mean SS (sd) | 108.6 (8.6) | 107.0 (8.4) | 107.3 (8.4) |

#### Criterion Validity

Within the research sample, word-finding difficulty was reported never/rarely (n=71, 46.7%), sometimes (n=43, 28.3%), or often (n=38, 25.0%); word-finding difficulty was more related to poorer performance on *Antonyms* than on other tasks, especially when adjusting for additional covariates (**Table 2**). Within the clinical sample, word-finding difficulty was reported as never/rarely (n=155, 30.5%), sometimes (n=172, 33.9%), or often (n=181, 35.6%); word-finding difficulty was most related to *Antonyms* (**Table 2**), which was the only task significantly related to patient-reported word-finding difficulty when adjusting for additional covariates. Within the combined sample (n=660), word-finding difficulty was reported as never/rarely (n=226, 34.2%), sometimes (n=215, 32.6%), or often (n=219, 33.2%). *Antonyms* was the only task that differed across patient-reported word-finding difficulty in the fully-adjusted model (**Table 2**).

**Table 2.**
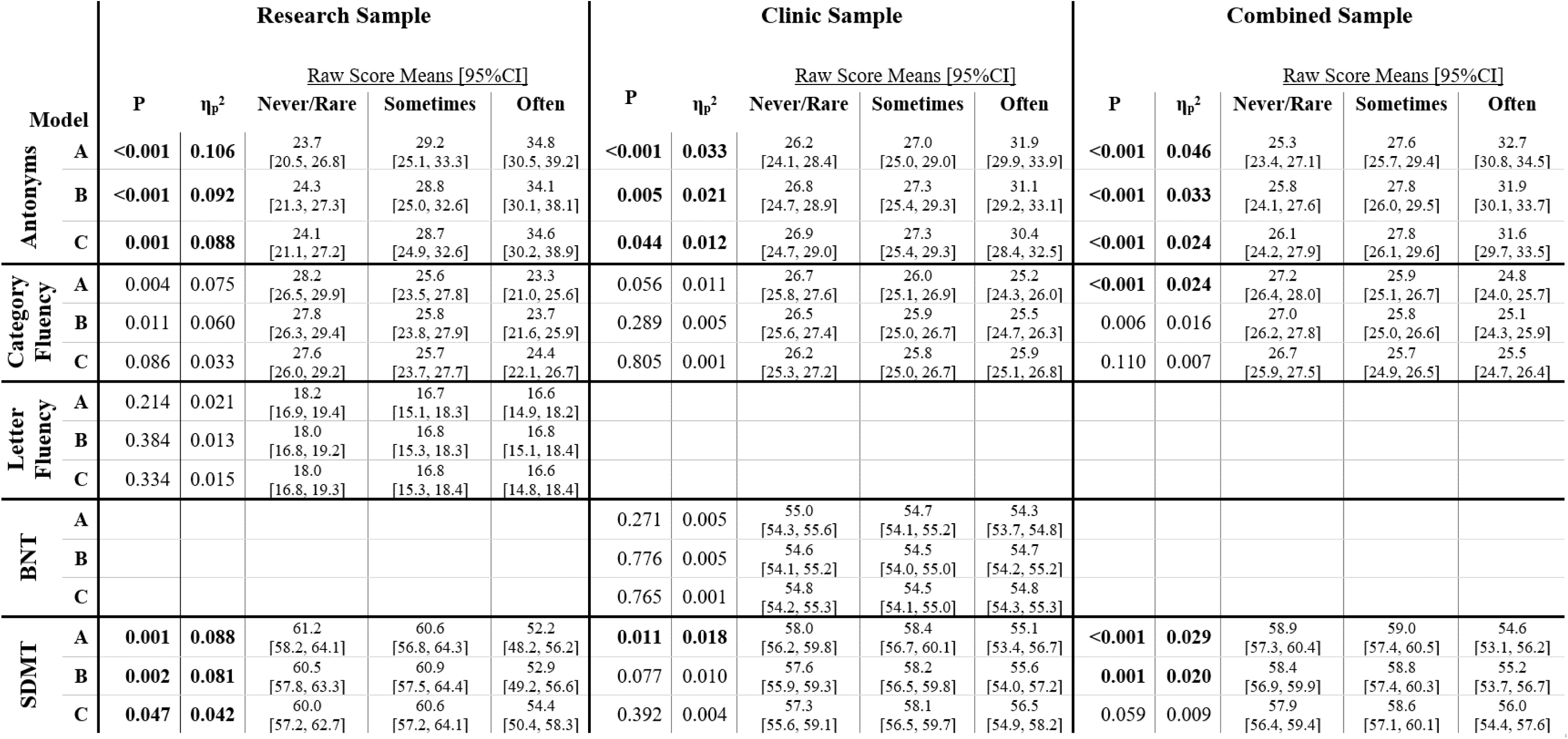
Task performance across levels of patient-reported word-finding difficulty. ANCOVAs investigated performance differences on each task (labeled on the left) and levels of patient-reported word-finding difficulty (never/rare, sometimes, often) adjusting for age, sex, education, and sample (Model A), additionally adjusting for premorbid verbal ability (Model B), and additionally adjusting for depression symptoms (Model C). Analyses were performed for separately for the research, clinical, and combined samples. *Antonyms* was the only task related to patient-reported word-finding across all models.

#### Discriminant Validity

*Antonyms* was significantly more related to MSCS Expressive Language than to Working Memory, Episodic Memory, or Executive/Speed (**Table 3**). Moreover, multiple regressions identified Expressive Language as the only MSCS subscale independently related to *Antonyms* performance (**Table S2**).

**Table 3.** Discriminant Validity. Partial correlations are shown between *Antonyms* performance and MSCS subscales adjusting for age, sex, education, and sample (A), additionally adjusting for premorbid verbal ability (B), and additionally adjusting for depression symptoms (C). Steiger Z tests indicated a stronger relationship between *Antonyms* performance and patient-reported difficulties in Expressive Language than between *Antonyms* performance and difficulties in Working Memory, Episodic Memory, or Executive/Speed.

| **Model** | **Expressive Language** | **Working Memory** | **Episodic Memory** | **Executive / Speed** |
| --- | --- | --- | --- | --- |
| **A** | .266 | .193<sup>*</sup> | .191<sup>*</sup> | .142<sup>**</sup> |
| **B** | .233 | .162<sup>*</sup> | .150<sup>*</sup> | .141<sup>*</sup> |
| **C** | .211 | .132<sup>*</sup> | .117<sup>*</sup> | .101<sup>**</sup> |

For each model (A, B, C), Steiger Z tests assessed whether the correlation between *Antonyms* and MSCS Expressive Language significantly differed from correlations between *Antonyms* and the other MSCS subscales; Bonferroni-adjusted P-values: <0.05<sup>\*</sup> <0.01<sup>\*\*</sup>

## DISCUSSION

*Antonyms* has good reliability and validity as a performance metric of word-finding difficulty in MS. Patient-reported word-finding difficulty was more related to performance on *Antonyms* than on traditional language tasks (Category Fluency, Letter Fluency, BNT) or general MS-related cognitive difficulty (SDMT). *Antonyms* also demonstrated discriminant validity as a metric specifically assessing word-finding / expressive language difficulty, rather than other cognitive functions.

Traditional rapid word generation tasks were initially developed to evaluate verbal fluency in the context of aphasia; these tasks appear less sensitive to MS-related word-finding difficulties, which are subtle but nonetheless impactful during early adulthood. Verbal fluency tasks require generation of *any* words within broad categories, unlike the naturalistic word-finding demand to rapidly retrieve *one specific* target word. Outside of aphasia, rapid word generation (verbal fluency) tasks are often interpreted as metrics of cognitive speed or executive function rather than language.^11–12^ Slowed cognitive speed may explain worse verbal fluency among persons with MS evaluated decades ago,^13^ leading to inclusion of verbal fluency within MS cognitive test batteries.^14–15^ Importantly, unlike historical MS cohorts, cognitive speed is normal among current patients diagnosed within the modern diagnostic and treatment era,^1^ for whom verbal fluency performance is also normal, but reported word-finding difficulty remains prevalent. Note that *Antonyms* does require rapid performance; however, subtraction of control task time isolates word-finding latency from general cognitive speed and articulation.

*Antonyms* was designed to assess phonological code retrieval by reducing semantic processing requirements through the use of strong word-pair associations. Implication of phonological code retrieval aligns with our observation that phonemic processing is below expectations in MS, and linked to worse patient-reported word-finding difficulty.^6^ A focus on phonological code retrieval is also consistent with radiologic evidence from ultra-high field MRI identifying the planum temporale / temporoparietal region as particularly vulnerable to cortical lesion formation in MS;^16^ phonological code retrieval is the chief language-related function of this region.^17^

Word-finding / phonological code retrieval needs further investigation, including whether MS-related word-finding difficulty stems from failure to activate specific codes and/or failure to inhibit competing codes (i.e., to retrieve “large,” one must inhibit synonyms such as “big,” “large,” and “huge”; see work on language disorders^18^). We need collaboration with experts in linguistics, neurophysiology and neuromodulation of language systems, and sub-millimeter neuroimaging of subtle perisylvian pathology. As a validated performance metric of MS-related word-finding difficulty, *Antonyms* will help to advance understanding and eventual treatment of this prevalent symptom of great importance to persons living with MS.

## Data Availability

All data produced in the present study are available upon reasonable request to the authors.

## ACKNOWLEDGEMENTS

We thank our colleagues at the Corinne Goldsmith Dickinson Center for Multiple Sclerosis at Mount Sinai Hospital, including the neurologists, neuropsychologists, and research coordinators. We also thank our research participants living with MS. This project was funded by the National Center for Medical Rehabilitation Research (NCMRR) within the National Institute of Child Health and Development (NICDH) of the National Institutes of Health (NIH; R01 HD082176).

## POTENTIAL CONFLICTS OF INTEREST

Nothing to report.

## ONLINE SUPPLEMENT

**Table S1.** Sample for 50-Item Pilot *Antonyms* Task. Analyses were completed with data from 170 native English speakers with relapsing-remitting multiple sclerosis (RRMS) enrolled within the Reserve against Disability in MS (RADIEMS) longitudinal cohort study, including 156 original participants completing their three-year follow-up, and 14 participants from a second wave of recruitment conducted at year three.

| Characteristic | Value |
| --- | --- |
| Age, years, mean (sd) | 37.7 (7.8) |
| Sex, N (%) |  |
| Women | 114 (67.1) |
| Men | 56 (32.9) |
| Race and Ethnicity, N (%) |  |
| Asian | 6 (3.5) |
| Black, non-Latino/a | 30 (17.6) |
| Latino/a | 37 (21.8) |
| White, non-Latino/a | 97 (57.1) |
| Bachelor's degree, N (%) | 133 (78) |
| Picture Vocabulary (NIH-TB), SS, mean (sd) | 105.2 (13.5) |
| EDSS, median (IQR) | 1.0 (0.0, 2.0) |
| Years since diagnosis, mean (sd) | 5.0 (3.9, 6.3) |

**Table S2.** Multiple regression predicting *Antonyms* with MSCS subscales. In the combined sample (n=660), multiple regressions predicted *Antonyms* performance with MSCS subscales in partially-adjusted and fully-adjusted models. In both models, Expressive Language was the only MSCS subscale independently predicting *Antonyms* performance.

| Predictor | Partially-Adjusted Model |  |  |  |  | Fully-Adjusted Model |  |  |  |  |
| --- | --- | --- | --- | --- | --- | --- | --- | --- | --- | --- |
| | B | 95% CI | | $\beta$ | p | B | 95% CI | | $\beta$ | p |
| Constant | 48.15 | 35.91 | 60.38 |  | <0.001 | 92.11 | 76.52 | 107.7 |  | <0.001 |
| Age | -0.12 | -0.23 | -0.01 | -0.08 | 0.041 | -0.08 | -0.18 | 0.03 | -0.05 | 0.146 |
| Sex | 0.30 | -2.15 | 2.75 | 0.01 | 0.811 | -0.07 | -2.39 | 2.26 | 0.00 | 0.956 |
| Education | -1.37 | -2.01 | -0.73 | -0.16 | <0.001 | -0.20 | -0.87 | 0.47 | -0.02 | 0.557 |
| Sample | 0.66 | -1.85 | 3.17 | 0.02 | 0.604 | 1.47 | -0.93 | 3.88 | 0.04 | 0.229 |
| Premorbid Verbal Ability |  |  |  |  |  | -0.59 | -0.73 | -0.46 | -0.34 | <0.001 |
| Depression Symptoms |  |  |  |  |  | 0.17 | -4.53 | 4.86 | 0.00 | 0.945 |
| MSCS Working Memory | -0.06 | -1.98 | 1.85 | 0.00 | 0.949 | -0.59 | -2.41 | 1.24 | -0.04 | 0.528 |
| MSCS Expressive Language | 3.53 | 2.04 | 5.01 | 0.28 | <0.001 | 3.13 | 1.72 | 4.55 | 0.25 | <0.001 |
| MSCS Episodic Memory | 0.68 | -0.91 | 2.27 | 0.05 | 0.402 | -0.03 | -1.55 | 1.49 | 0.00 | 0.973 |
| MSCS Executive / Speed | -0.77 | -2.11 | 0.57 | -0.06 | 0.261 | 0.16 | -1.23 | 1.56 | 0.01 | 0.817 |

**Figure S1.**
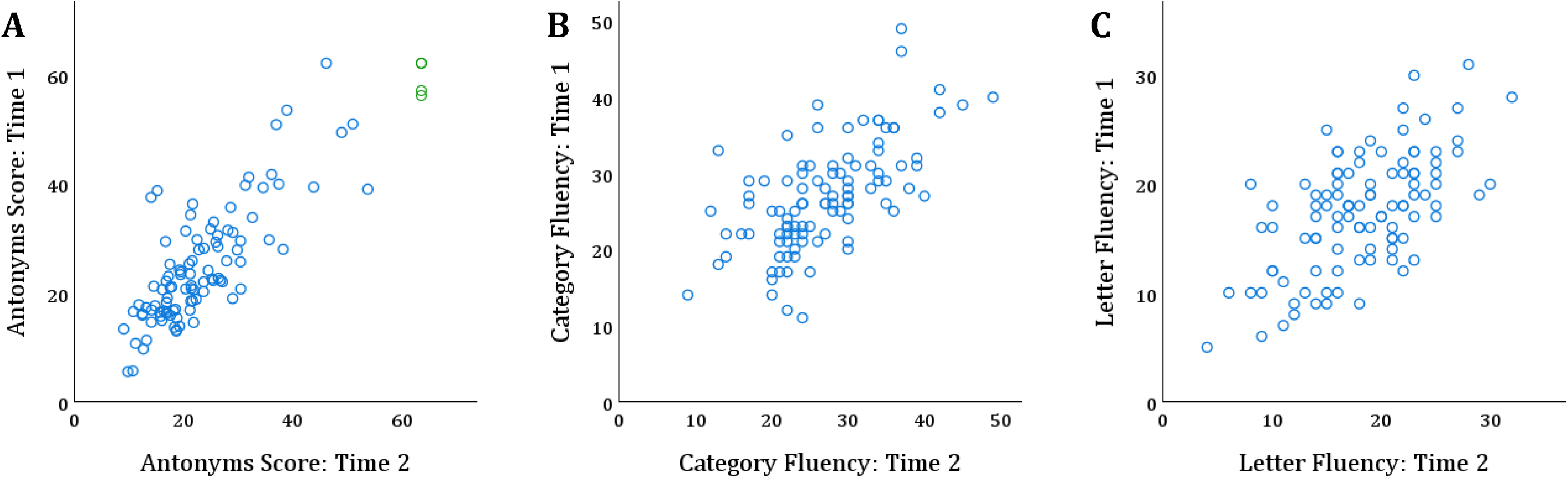
Scatterplots compare Time 1 and Time 2 scores for *Antonyms* (panel A), Category Fluency (panel B), and Letter Fluency (panel C) over a median (IQR) interval of 23.7 (18.5, 26.0) months in a sample of 102 patients with relapse-onset MS (mean [sd] age of 41.7 [7.5] years; 64.7% female; 61.8% non-Hispanic White; diagnosed median [IQR] of 8.3 [7.2, 9.6] years). Scores at Time 1 and Time 2 did not differ within patients for any task (Ps>0.05). Reliability (ICC [95%CI]; two-way mixed, absolute agreement) of *Antonyms* was good (0.85[0.79, 0.90]) and better than reliabilities of traditional verbal fluency tasks requiring rapid generation of animals (Category Fluency; 0.64[0.51, 0.74]) or words starting with “f” (Letter Fluency; 0.61[0.48, 0.72]). In panel A, green circles represent four patients (two overlapping) with extreme scores (>3.0*IQR) that were winsorized; however, reliability of *Antonyms* was nearly identical with (0.85[0.79, 0.90]) without (0.86[0.80, 0.90]) winsorizing. There were no extreme scores for Category Fluency or Letter Fluency. [Test-retest reliability was re-examined within a subsample of 15 patients reassessed after a shorter interval of ≥1 month but <6 months (3.0 [1.8, 3.7] months); despite the small sample, test-retest reliability was still good for *Antonyms* (0.84 [0.60, 0.94]), and still better than for Category Fluency (0.33 [-0.18, 0.70]) and Letter Fluency (0.42 [-0.12, 0.76]).]

## APPENDIX

### Antonyms Task

The following pages include:

- Antonyms Record Form
- “State the Opposite” stimulus page with instructions and practice items
- Rapid Antonym Retrieval stimulus test page
- “Read the Word Pairs” stimulus page with instructions and practice items
- Rapid Word-Pair Reading stimulus test page

### Antonyms Record Form

#### Rapid Antonym Retrieval

##### Instructions

Present “State the Opposite” page, read directions and sample, then have person perform practice items. Make sure person reads cue words aloud. Read directions for “Test Items,” then present the test page, emphasize that person should work as quickly as possible down the first column then immediately continue with the second column. Say “Ready? Go!”

If person pauses on an item for four seconds, instruct them to move on to the next item.

Record responses that differ from those listed below; alternate correct responses are provided in parentheses, but consult a thesaurus if you believe a different response may also be correct. (Note, however, that completion time is the primary outcome; accuracy is secondary.)

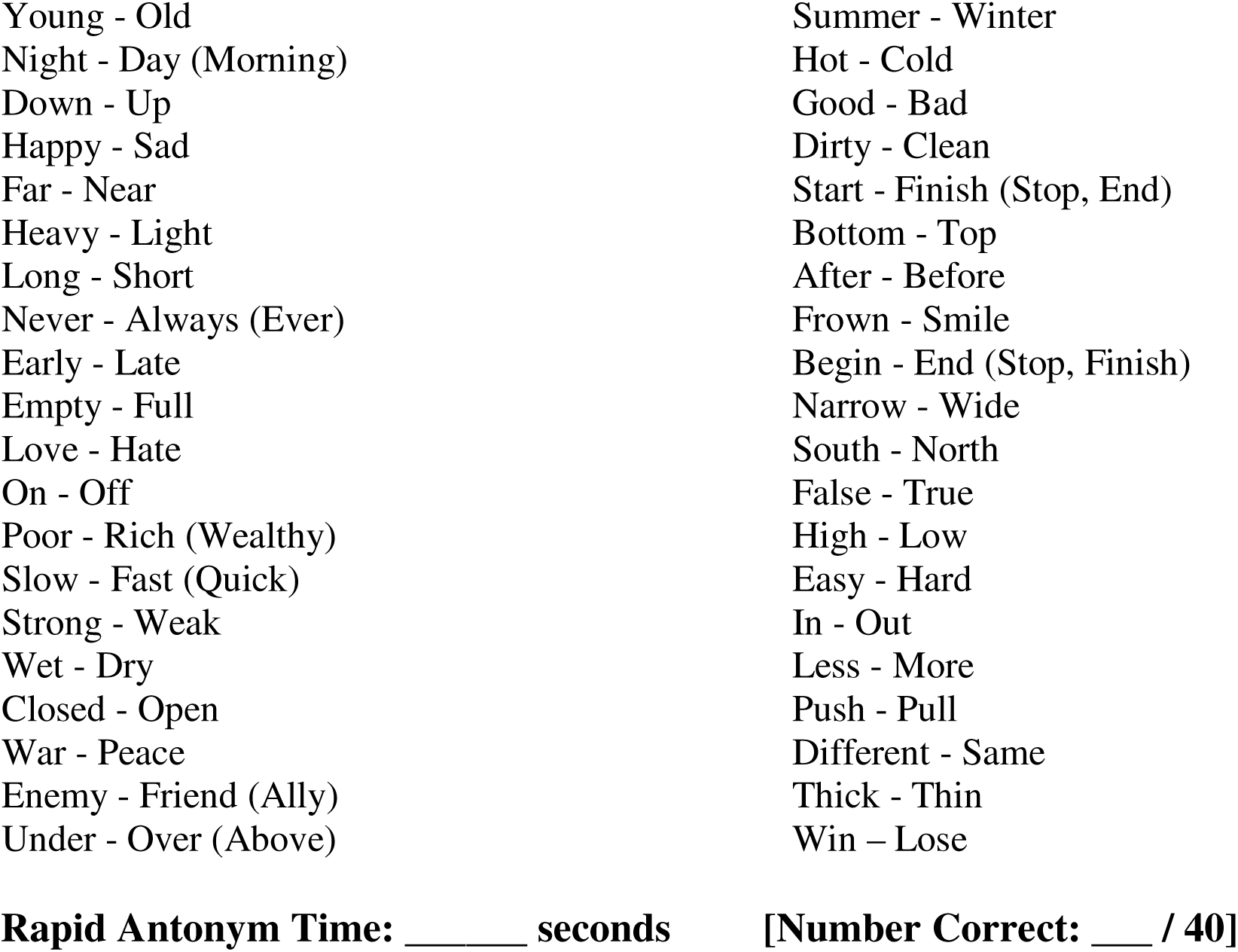

#### Rapid Word-Pair Reading (Control Task)

##### Instructions

Present “Read the Word Pairs” page, read directions and sample, then have person perform practice items. Read directions for “Test Items,” then present the test page, emphasize that person should work as quickly as possible down the first column then immediately continue with the second column. Say “Ready? Go!”

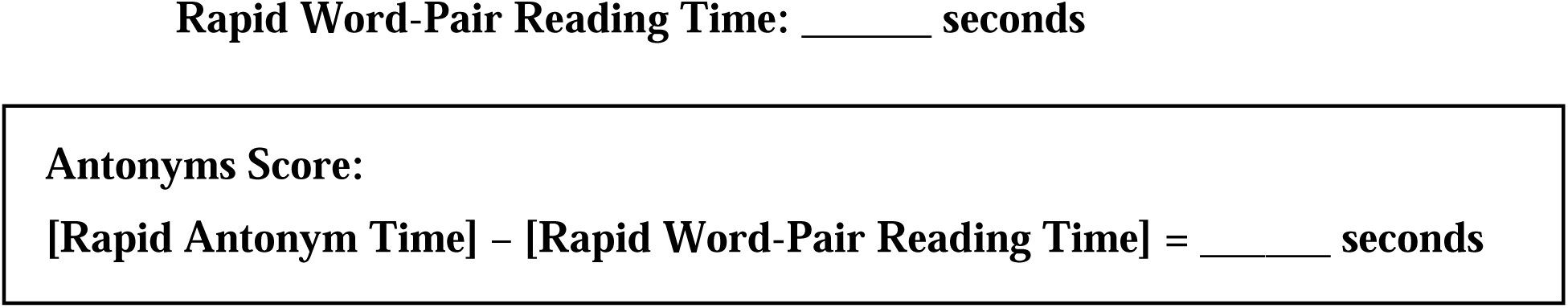

### State the Opposite

#### Directions

Quickly read each word aloud and state the opposite of that word aloud.

Sample: If you saw “**Yes - ____**” you would say “**Yes – No**.”

#### Practice

Quickly read each word aloud and state the opposite of that word. Keep going as quickly as possible until you finish the last word.

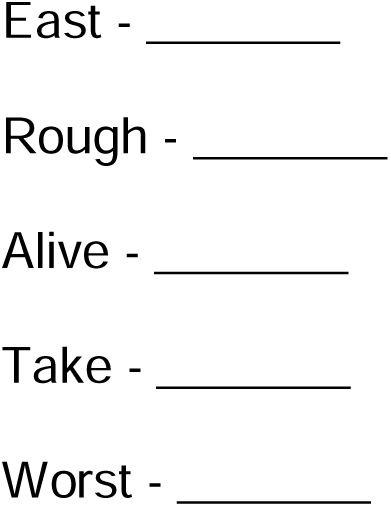

#### Test Items

Next you will see two columns of words. Quickly complete the first column and then complete the second column. Work as quickly as you can until the end.

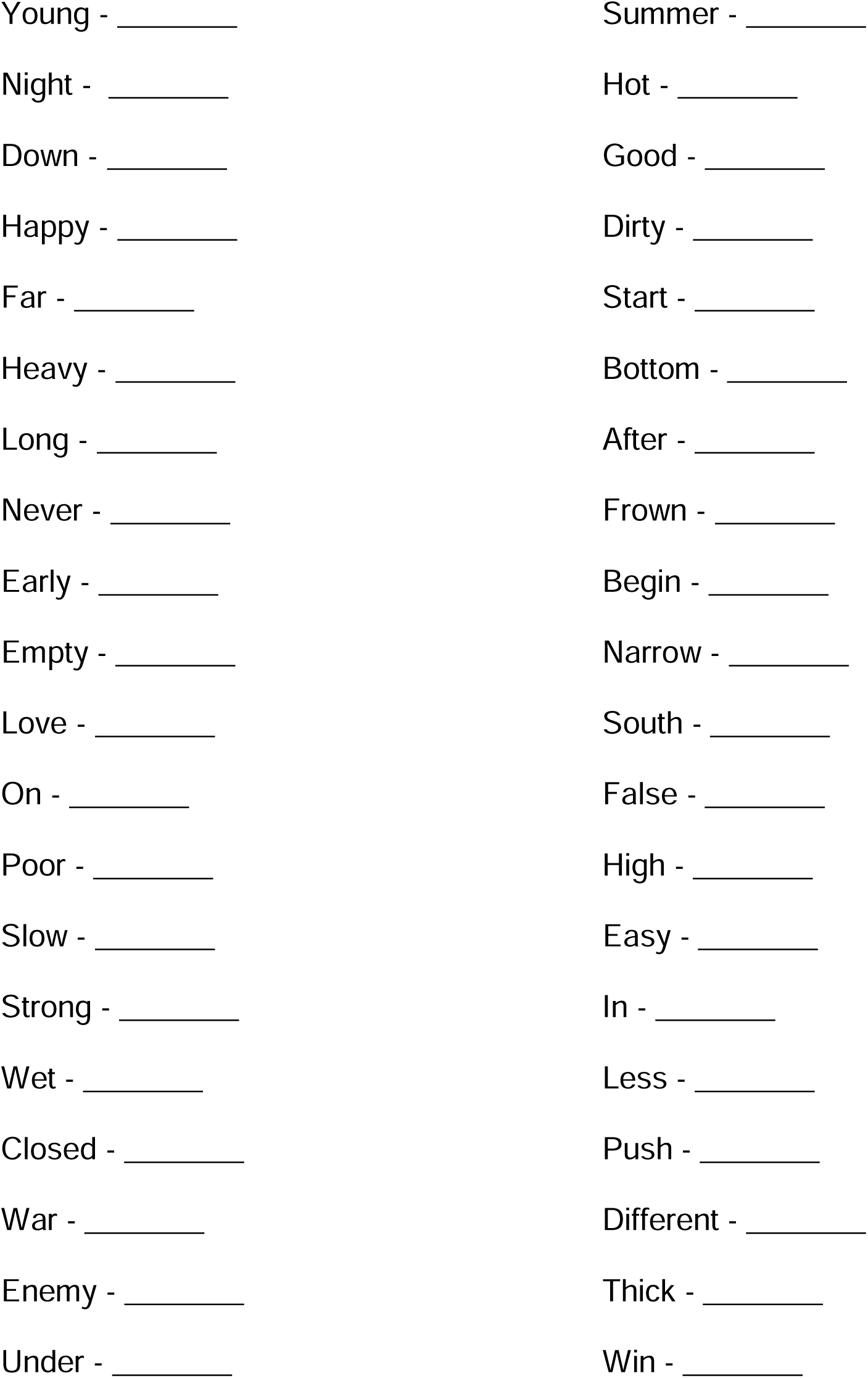

### Read the Word Pairs

#### Directions & Practice

Quickly read each word pair aloud. For instance, if you saw “**Yes - No**” you would say “**Yes - No**” aloud.

Read the following word pairs as quickly as possible until you finish the last pair.

East - West
Rough - Smooth
Alive - Dead
Take - Give
Worst - Best

#### Test Items

Next you will see two columns of words pairs. Quickly read the first column aloud and then the second column. Some answers may be different from what you said before, but just read what is printed on the page. Work as quickly as you can until the end.

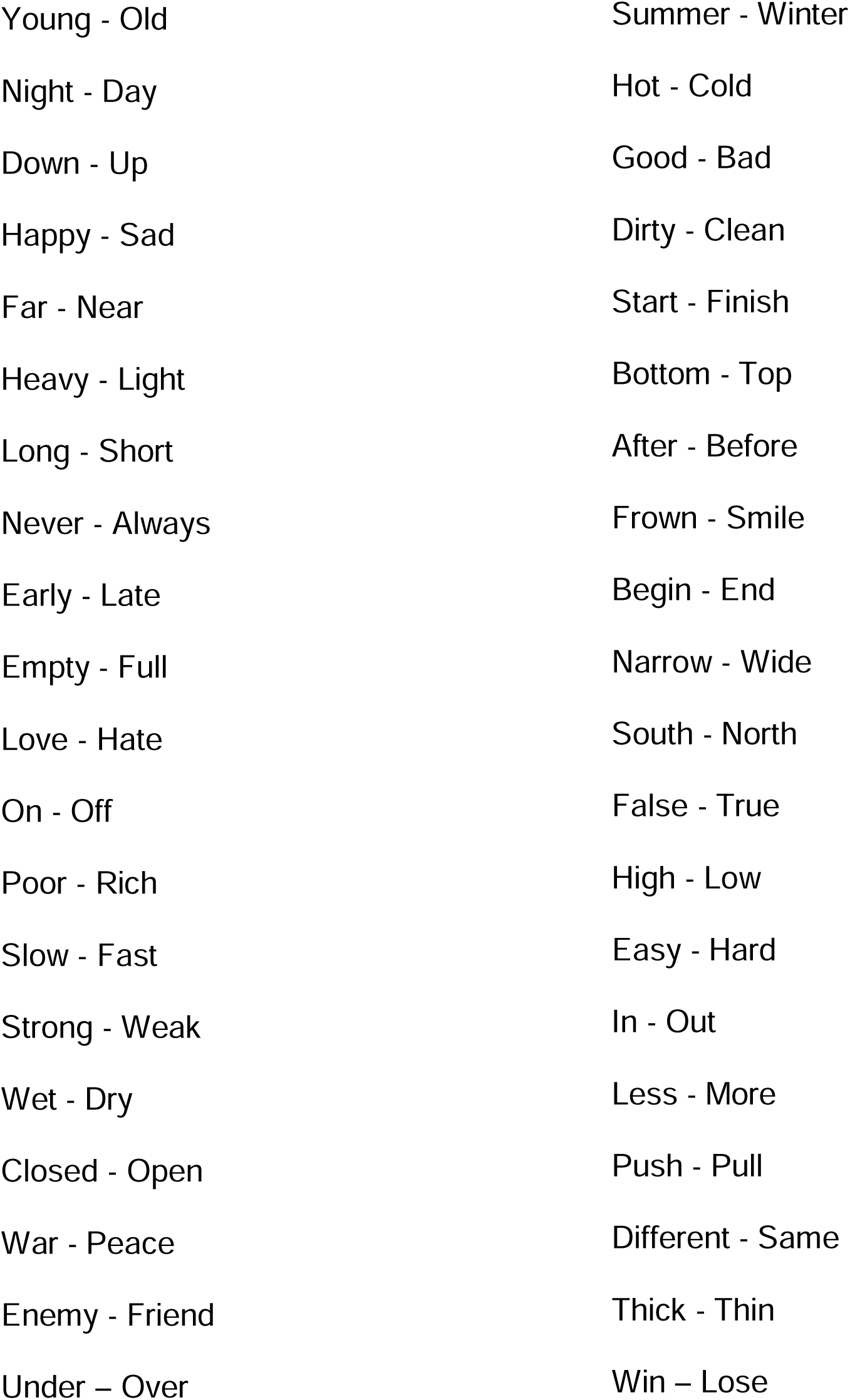

## Notes

### Competing Interest Statement

The authors have declared no competing interest.

### Author Declarations

Ethic committee/IRB of the Icahn School of Medicine at Mount Sinai gave ethical approval for this work.

